# Association of Toothbrushing, Dental Flossing, and Interdental Brushing with Stroke Risk

**DOI:** 10.1101/2025.08.04.25332999

**Authors:** Sangwoo Park, Da Eun Kim, Sun Jae Park, Jihun Song, Hye Jun Kim, Su Kyoung Lee, Hyun-Young Shin, Hyun-Jae Cho, Sang Min Park

**Affiliations:** Department of Biomedical Sciences, Seoul National University Graduate School, Seoul, Republic of Korea; Institute of Health and Environment Graduate School of Public Health Seoul National University, Seoul, Republic of Korea; Department of Family Medicine, Seoul National University Hospital, Seoul, Republic of Korea; Department of Family Medicine, Seoul St. Mary’s Hospital, Seoul, Republic of Korea; Department of Preventive and Public Oral Health, School of Dentistry, Seoul National University, Seoul, Republic of Korea

## Abstract

**Background:** Stroke is the second leading cause of death globally. While daily toothbrushing is widely promoted for oral hygiene, the preventive impact of adjunctive practices such as dental flossing and interdental brushing on stroke remains unclear.

**Objectives:** This study aimed to investigate the association between comprehensive oral hygiene behaviors and the risk of stroke.

**Methods:** We conducted a population-based retrospective cohort study using data from the National Health Insurance Service–Health Screening cohort in Korea. A total of 98,866 adults aged 40 years or older who underwent both general and oral health examinations during 2009–2010 were included. Participants were followed from January 2011 to December 2019. Individuals with pre-existing cardiovascular disease, death before baseline, or missing data were excluded. Oral hygiene practices, including daily toothbrushing frequency and weekly use of dental floss and interdental brushes, were self-reported. The primary outcome was stroke requiring hospitalization for ≥2 days, classified as total, ischemic, or hemorrhagic stroke based on diagnostic codes. Multivariable Cox regression models were used to estimate adjusted hazard ratios.

**Results:** Compared to individuals with poor oral hygiene, participants who brushed their teeth at least twice daily and consistently used dental floss and interdental brushes showed a 23% lower risk of ischemic stroke. A significant trend was observed across oral hygiene behavior categories.

**Conclusions:** Frequent toothbrushing, along with regular use of dental floss and interdental brushes, was associated with a reduced risk of ischemic stroke. Promoting comprehensive oral hygiene may offer additional benefits in stroke prevention.

**Key Points:** *Question:* Is improved oral hygiene practices, including toothbrushing, dental flossing, and interdental brushing, associated with a reduced risk of stroke?

*Findings:* In this nationwide retrospective cohort study that included 98,866 participants, more frequent toothbrushing, particularly dental flossing, and interdental brushing were significantly associated with up to a 23% reduction in the risk of ischemic stroke, compared to those in the poorest oral health care group.

*Meaning:* Enhancing oral hygiene practices such as toothbrushing, dental flossing and interdental brushing may significantly reduce the risk of stroke.

## 1. Introduction

Stroke ranks as the world’s second leading cause of mortality among adults^1^. High stroke severity can negatively impact the quality of life^2^. Various modifiable risk factors for stroke have been well-documented^3^. Notably, poor oral hygiene, including dental caries^4^, tooth loss^5–7^, and periodontitis^8–10^, is also considered as a potential contributor. This suggests that enhancing oral health could potentially lower the risk of stroke. Given that recent research indicates that up to 85% of all strokes may be avoidable^11^, it is essential to manage stroke risk factors early, before a stroke occurs.

Effective oral hygiene practices include regular toothbrushing and the use of interdental cleaning methods^12,13^. Several studies have established the link between toothbrushing and cardiovascular diseases. However, little research has specifically investigated the impact of dental flossing and interdental brushing on stroke risk beyond the importance of toothbrushing. Toothbrushing alone is insufficient for removing biofilms in interdental areas due to its limited reach^14,15^. In contrast, interdental cleaners can more effectively reach these spaces and remove plaque^15^. Therefore, using interdental cleaning devices is crucial for addressing the limitations of toothbrushing and enhance oral hygiene^16^.

Our study was motivated by the potential impact of dental floss and interdental toothbrushes on stroke risk. We aimed to provide novel insights into how the frequency of these oral hygiene practices might influence stroke risk, utilizing data from the National Health Insurance Service (NHIS) of the Republic of Korea.

## 2. Methods

Data were retrieved from the NHIS of the Republic of Korea, which provides compulsory healthcare coverage to approximately 97% of the population^17^. South Koreans aged 20 and above are eligible for biannual health screening programs, which include anthropometric measurements, laboratory findings, and self-reported questionnaires for lifestyle activities^18^. The NHIS also offers a dental screening program, comprising professional dental checkups and self-reported questionnaires on oral hygiene care^19^. Hence, the NHIS database includes sociodemographic details, health and dental screening results, hospital visit records, and prescription records^17^. The NHIS database has been validated elsewhere^17^.

This study included 130,789 participants who completed health and dental screening programs of the NHIS between January 1, 2009, and December 31, 2010. The index date was designated as January 1, 2011. We excluded participants with diagnosed cardiovascular disease (n=15,542) or those who passed away (n=248) before the index date, those with missing data for toothbrushing (n=2,091), those with missing data for dental flossing and interdental brushing or those who were unaware of these practices (n=9,226), and those with missing covariate values (n=4,816). Consequently, a total of 98,866 participants were monitored from January 1, 2011, until the first occurrence of a newly diagnosed stroke, death, or December 31, 2019 (Figure 1).

**Figure 1.**
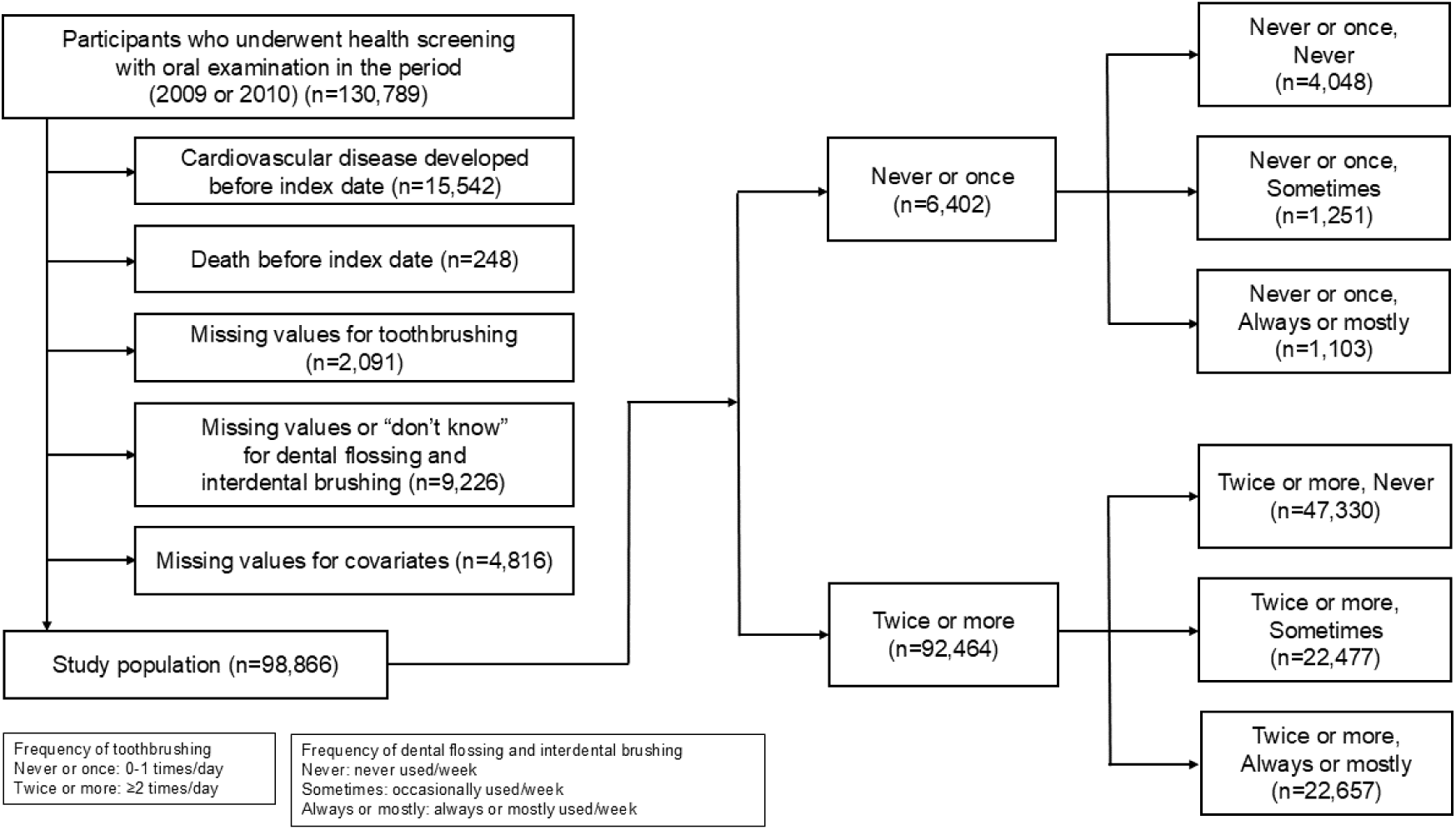
Flow diagram of the study population selection process.

The frequency of toothbrushing, dental flossing, and interdental brushing was estimated through self-reported questionnaires completed by participants during the oral examination. First, we classified participants into two groups based on their daily toothbrushing frequency: a ‘never or once’ group (0 to 1 times) and a ‘twice or more’ group (2 or more times^20^). Next, these two groups were further classified into three subgroups according to their weekly frequency of dental flossing and interdental brushing: a ‘never’ group (never used), a ‘sometimes’ group (occasionally used), and an ‘always or mostly’ group (always or mostly used). Thus, there were a total of six groups in accordance with toothbrushing frequency and the frequency of dental flossing and interdental brushing.

The outcome variable was the incidence of stroke, defined as hospitalization for 2 days or more with ICD-10 codes (total stroke: I60-I69; ischemic stroke: I63; and hemorrhagic stroke: I60-I62), which were also used to evaluate the onset of stroke in Korea^21–23^.

The covariates included for analysis were age, sex, body mass index (BMI), systolic blood pressure, diastolic blood pressure, fasting serum glucose, total cholesterol, household income, smoking status, alcohol consumption, physical activity, missing teeth, and Charlson comorbidity index (CCI). Smoking status, alcohol consumption, physical activity, and missing teeth were assessed through self-reported questionnaires. The CCI was calculated in accordance with a previous study^24^.

Baseline characteristics of the study population are described in Table 1. We carried out multivariable Cox proportional hazards regression analyses utilizing two models to evaluate the impact of oral hygiene practices on stroke risk. The first model was adjusted for age and sex, while the second model was additionally adjusted for all other covariates. Adjusted hazard ratios (aHRs) and 95% confidence intervals (CIs) were calculated for stroke risk. Furthermore, we carried out a stratified analysis for stroke risk within subgroups of age, sex, household income, smoking status, alcohol consumption, physical activity, missing teeth, and the CCI. All statistical analyses were conducted utilizing SAS Enterprise Guide 7.1 (SAS Institute Inc.), with a two-sided p-value threshold of <0.05 used to determine statistical significance.

**Table 1.** Descriptive characteristics of the study population.

| Frequency of toothbrushing (times/day) | Total | Never or once |  |  | Twice or more |  |  |
| --- | --- | --- | --- | --- | --- | --- | --- |
| Frequency of dental flossing and interdental brushing (frequency/week) |  | Never | Sometimes | Always or mostly | Never | Sometimes | Always or mostly |
| Number of participants (%) | 98,866 | 4,048 (4.09) | 1,251 (1.27) | 1,103 (1.12) | 47,330 (47.87) | 22,477 (22.73) | 22,657 (22.92) |
| Age, years, mean (SD) | 56.01 (7.42) | 60.30 (8.95) | 57.45 (7.45) | 57.76 (7.60) | 56.25 (7.69) | 54.69 (6.38) | 55.89 (7.14) |
| Sex, N (%) |  |  |  |  |  |  |  |
| Male | 58,725 (59.40) | 2,966 (73.27) | 998 (79.78) | 801 (72.62) | 26,435 (55.85) | 13,702 (60.96) | 13,823 (61.01) |
| Female | 40,141 (40.60) | 1,082 (26.73) | 253 (20.22) | 302 (27.38) | 20,895 (44.15) | 8,775 (39.04) | 8,834 (38.99) |
| Body mass index, kg/m<sup>2</sup>, mean (SD) | 23.92 (2.81) | 24.11 (2.99) | 24.13 (3.01) | 23.98 (2.93) | 23.95 (2.82) | 23.91 (2.76) | 23.83 (2.79) |
| Systolic blood pressure, mmHg, mean (SD) | 123.84 (14.62) | 125.38 (15.24) | 125.32 (15.04) | 124.55 (14.57) | 123.88 (14.70) | 123.34 (14.38) | 123.86 (14.54) |
| Diastolic blood pressure, mmHg, mean (SD) | 77.25 (9.79) | 77.68 (9.86) | 78.13 (10.04) | 77.65 (9.89) | 77.17 (9.82) | 77.19 (9.71) | 77.32 (9.78) |
| Fasting serum glucose, mg/dL, mean (SD) | 99.95 (23.75) | 102.94 (27.25) | 104.71 (30.83) | 103.77 (29.60) | 99.80 (23.07) | 99.42 (23.04) | 99.82 (24.33) |
| Total cholesterol, mg/dL, mean (SD) | 200.73 (35.94) | 197.86 (36.90) | 198.69 (35.63) | 200.38 (37.74) | 200.88 (36.00) | 201.05 (35.60) | 200.72 (35.88) |
| Household income<sup>a</sup>, quartiles, N (%) |  |  |  |  |  |  |  |
| 1st (highest) | 41,011 (41.48) | 1,318 (32.56) | 449 (35.89) | 375 (34.00) | 19,156 (40.47) | 10,397 (46.26) | 9,316 (41.12) |
| 2nd | 19,692 (19.92) | 940 (23.22) | 269 (21.50) | 229 (20.76) | 9,317 (19.69) | 4,319 (19.22) | 4,618 (20.38) |
| 3rd | 20,183 (20.41) | 919 (22.70) | 283 (22.62) | 259 (23.48) | 9,924 (20.97) | 4,207 (18.72) | 4,591 (20.26) |
| 4th (lowest) | 17,980 (18.19) | 871 (21.52) | 250 (19.98) | 240 (21.76) | 8,933 (18.87) | 3,554 (15.81) | 4,132 (18.24) |
| Smoking status, N (%) |  |  |  |  |  |  |  |
| Never smoker | 59,020 (59.70) | 1,989 (49.14) | 524 (41.89) | 596 (54.03) | 29,219 (61.73) | 13,022 (57.93) | 13,670 (60.33) |
| Past smoker | 21,347 (21.59) | 994 (24.56) | 388 (31.02) | 253 (22.94) | 9,562 (20.20) | 5,263 (23.42) | 4,887 (21.57) |
| Current smoker | 18,499 (18.71) | 1,065 (26.31) | 339 (27.10) | 254 (23.03) | 8,549 (18.06) | 4,192 (18.65) | 4,100 (18.10) |
| Alcohol consumption, N (%) |  |  |  |  |  |  |  |
| None | 53,475 (54.09) | 1,997 (49.33) | 521 (41.65) | 584 (52.95) | 26,561 (56.12) | 11,663 (51.89) | 12,149 (53.62) |
| 2-3 times/month | 31,450 (31.81) | 1,127 (27.84) | 437 (34.93) | 326 (29.56) | 14,361 (30.34) | 7,756 (34.51) | 7,443 (32.85) |
| 1-2 times/week | 9,915 (10.03) | 509 (12.57) | 169 (13.51) | 127 (11.51) | 4,555 (9.62) | 2,335 (10.39) | 2,220 (9.80) |
| ≥3 times/week | 4,026 (4.07) | 415 (10.25) | 124 (9.91) | 66 (5.98) | 1,853 (3.92) | 723 (3.22) | 845 (3.73) |
| Physical activity, times per week, N (%) |  |  |  |  |  |  |  |
| 0 | 41,109 (41.58) | 2,326 (57.46) | 546 (43.65) | 522 (47.33) | 21,189 (44.77) | 8,167 (36.33) | 8,359 (36.89) |
| 1-2 | 18,816 (19.03) | 639 (15.79) | 260 (20.78) | 180 (16.32) | 8,898 (18.80) | 4,557 (20.27) | 4,282 (18.90) |
| 3-4 | 15,854 (16.04) | 423 (10.45) | 187 (14.95) | 148 (13.42) | 7,043 (14.88) | 4,141 (18.42) | 3,912 (17.27) |
| ≥5 | 23,087 (23.35) | 660 (16.30) | 258 (20.62) | 253 (22.94) | 10,200 (21.55) | 5,612 (24.97) | 6,104 (26.94) |
| Missing teeth, N (%) |  |  |  |  |  |  |  |
| No | 73,473 (74.32) | 2,698 (66.65) | 835 (66.75) | 793 (71.89) | 34,705 (73.33) | 17,097 (76.06) | 17,345 (76.55) |
| Yes | 25,373 (25.68) | 1,350 (33.35) | 416 (33.25) | 310 (28.11) | 12,625 (26.67) | 5,380 (23.94) | 5,312 (23.45) |
| Charlson comorbidity index, N (%) |  |  |  |  |  |  |  |
| 0 | 15,858 | 545 | 210 | 176 | 7,535 | 3,859 | 3,533 |
|  | (16.04) | (13.46) | (16.79) | (15.96) | (15.92) | (17.17) | (15.59) |
| 1 | 23,267 | 822 | 263 | 217 | 11,029 | 5,516 | 5,420 |
|  | (23.53) | (20.31) | (21.02) | (19.67) | (23.30) | (24.54) | (23.92) |
| ≥2 | 59,741 | 2,681 | 778 | 710 | 28,766 | 13,102 | 13,704 |
|  | (60.43) | (66.23) | (62.19) | (64.37) | (60.78) | (58.29) | (60.48) |

Never or once: brushed teeth 0-1 times per day. Twice or more: brushed teeth 2 or more times per day
Never: never used dental floss and interdental brushes per week. Sometimes: occasionally used dental floss and interdental brushes per week. Always or mostly: always or mostly used dental floss and interdental brushes per week
Abbreviation: N= number of subjects; SD= standard deviation
Data are presented as median (interquartile range) unless otherwise specified.
<sup>a</sup>Proxy for socioeconomic status based on the insurance premium of the National Health Insurance Service

## 3. Results

Table 1 displays the initial characteristics of the study cohort. Among the 98,866 participants, 6,402 (6.5%) brushed their teeth 0 to 1 times daily, while 92,464 (93.5%) brushed their teeth at least twice daily. In terms of the frequency of weekly dental flossing and interdental brushing, 51,378 individuals (52.0%) never used dental floss or interdental brushes, 23,728 (24.0%) sometimes used them, and 23,760 (24.0%) always or mostly used them. Among all participants, 4,048 individuals (4.1%) who brushed their teeth 0 to 1 times daily and never used dental floss or interdental brushes weekly belonged to the poorest oral health care group. In contrast, 22,657 individuals (22.9%) who brushed their teeth at least twice daily and always or mostly used dental floss and interdental brushes weekly belonged to the highest oral health care group.

Table 2 presents the impact of the frequency of dental flossing and interdental brushing on stroke risk, both with and without considering the toothbrushing frequency. The poorest oral health care group served as the primary reference group, while another reference group included individuals who brushed their teeth at least twice daily but never used dental floss and interdental brushes weekly. A general trend toward reduced stroke risk was observed with increased toothbrushing frequency to at least twice daily compared to the poorest oral health care group. Similarly, an increased frequency of weekly dental flossing and interdental brushing was associated with a lower stroke risk, regardless of daily toothbrushing frequency. Notably, a significantly lower risk of ischemic stroke was observed in the group that brushed their teeth at least twice daily and always or mostly used dental floss and interdental brushes weekly, compared to both the poorest oral health care group (aHR 0.77, 95% CI 0.63–0.94) and the other reference group (aHR 0.88, 95% CI 0.79–1.00), according to the adjusted multivariable Cox proportional hazards model (*P*_trend_=0.008).

**Table 2.** Hazard ratios analysis of risk for stroke^a^ according to the frequency of toothbrushing, dental flossing and interdental brushing.

| Frequency of toothbrushing (times/day) | Never or once |  |  | Twice or more |  |  | p for trend |
| --- | --- | --- | --- | --- | --- | --- | --- |
| Frequency of dental flossing and interdental brushing (frequency/week) | Never | Sometimes | Always or mostly | Never | Sometimes | Always or mostly |  |
| Number of participants (%) | 4,048<br>(4.09) | 1,251<br>(1.27) | 1,103<br>(1.12) | 47,330<br>(47.87) | 22,477<br>(22.73) | 22,657<br>(22.92) |  |
| Person-years | 1,911 | 573 | 550 | 15,455 | 6,409 | 7,168 |  |
| **Total stroke<sup>a</sup>** |  |  |  |  |  |  |  |
| Number of cases | 256 | 71 | 59 | 1,965 | 749 | 853 |  |
| aHR (95% CI)<sup>b</sup> | **1.00**<br>(Ref.) | 1.02<br>(0.79-1.33) | 0.81<br>(0.61-1.07) | 0.97<br>(0.85-1.10) | 0.94<br>(0.81-1.08) | 0.91<br>(0.79-1.05) | 0.139 |
| aHR (95% CI)<sup>c</sup> | **1.00**<br>(Ref.) | 1.01<br>(0.77-1.33) | 0.81<br>(0.61-1.08) | 0.97<br>(0.84-1.11) | 0.94<br>(0.82-1.09) | 0.92<br>(0.79-1.06) | 0.202 |
| aHR (95% CI)<sup>b</sup> | **1.00**<br>(Ref.) | 1.02<br>(0.79-1.33) | 0.81<br>(0.61-1.07) | **1.00**<br>(Ref.) | 0.97<br>(0.89-1.05) | 0.94<br>(0.87-1.02) | 0.139 |
| aHR (95% CI)<sup>c</sup> | **1.00**<br>(Ref.) | 1.01<br>(0.77-1.33) | 0.81<br>(0.61-1.08) | **1.00**<br>(Ref.) | 0.98<br>(0.90-1.07) | 0.95<br>(0.88-1.03) | 0.202 |
| **Ischemic stroke<sup>d</sup>** |  |  |  |  |  |  |  |
| Number of cases | 148 | 39 | 35 | 881 | 328 | 365 |  |
| aHR (95% CI)<sup>b</sup> | **1.00**<br>(Ref.) | 1.00<br>(0.71-1.43) | 0.87<br>(0.60-1.26) | 0.86<br>(0.73-1.03) | 0.84<br>(0.69-1.02) | **0.76**<br>(0.63-0.92) | 0.002 |
| aHR (95% CI)<sup>c</sup> | **1.00**<br>(Ref.) | 1.00<br>(0.70-1.44) | 0.89<br>(0.61-1.29) | 0.88<br>(0.73-1.05) | 0.86<br>(0.70-1.05) | **0.77**<br>(0.63-0.94) | 0.008 |
| aHR (95% CI)<sup>b</sup> | **1.00**<br>(Ref.) | 1.00<br>(0.71-1.43) | 0.87<br>(0.60-1.26) | **1.00**<br>(Ref.) | 0.97<br>(0.85-1.10) | **0.88**<br>(0.78-1.00) | 0.002 |
| aHR (95% CI)<sup>c</sup> | **1.00**<br>(Ref.) | 1.00<br>(0.70-1.44) | 0.89<br>(0.61-1.29) | **1.00**<br>(Ref.) | 0.98<br>(0.86-1.12) | **0.88**<br>(0.79-1.00) | 0.008 |
| **Hemorrhagic stroke<sup>e</sup>** |  |  |  |  |  |  |  |
| Number of cases | 35 | 13 | 6 | 277 | 97 | 110 |  |
| aHR (95% CI)<sup>b</sup> | **1.00**<br>(Ref.) | 1.41<br>(0.74-2.67) | 0.62<br>(0.26-1.47) | 1.04<br>(0.72-1.49) | 0.92<br>(0.62-1.37) | 0.89<br>(0.61-1.32) | 0.243 |
| aHR (95% CI)<sup>c</sup> | **1.00**<br>(Ref.) | 1.37<br>(0.72-2.61) | 0.63<br>(0.26-1.51) | 0.98<br>(0.68-1.40) | 0.88<br>(0.59-1.32) | 0.85<br>(0.58-1.26) | 0.256 |
| aHR (95% CI)<sup>b</sup> | **1.00**<br>(Ref.) | 1.41<br>(0.74-2.67) | 0.62<br>(0.26-1.47) | **1.00**<br>(Ref.) | 0.89<br>(0.70-1.12) | 0.86<br>(0.69-1.07) | 0.243 |
| aHR (95% CI)<sup>c</sup> | **1.00**<br>(Ref.) | 1.37<br>(0.72-2.61) | 0.63<br>(0.26-1.51) | **1.00**<br>(Ref.) | 0.90<br>(0.71-1.14) | 0.87<br>(0.70-1.09) | 0.256 |

Never or once: brushed teeth 0-1 times per day. Twice or more: brushed teeth 2 or more times per day
Never: never used dental floss and interdental brushes per week. Sometimes: occasionally used dental floss and interdental brushes per week. Always or mostly: always or mostly used dental floss and interdental brushes per week
Abbreviation: aHR= adjusted hazard ratio; CI= confidence interval
<sup>a</sup>Defined as subjects who had experienced at least two days of hospitalization with I60–I69
<sup>b</sup>Cox proportional hazard model adjusted for age and sex.
<sup>c</sup>Cox proportional hazard model adjusted for age, sex, body mass index, systolic blood pressure, diastolic blood pressure, fasting serum glucose, total cholesterol, household income, smoking status, alcohol consumption, physical activity, missing teeth, and the Charlson Comorbidity Index.
<sup>d</sup>Defined as subjects who had experienced at least two days of hospitalization with I63
<sup>e</sup>Defined as subjects who had experienced at least two days of hospitalization with I60-I62

Table 3 shows the results of the stratified analysis for total stroke risk by various covariates, such as age, sex, household income, smoking status, alcohol consumption, physical activity, missing teeth, and the CCI, with the poorest oral health care group as the reference. Compared to this group, a statistically significant decrease in total stroke risk was observed among individuals under 65 years of age (aHR 0.78, 95% CI 0.65–0.95), those who have ever smoked (aHR 0.79, 95% CI 0.64–0.97), and those with missing teeth (aHR 0.77, 95% CI 0.60–0.99), when they brushed their teeth at least twice daily and always or mostly used dental floss and interdental brushes weekly (all *P*_trend_<0.05). Furthermore, we observed a significantly lower risk of total stroke among individuals with missing teeth (aHR 0.59, 95% CI 0.36–0.98), when these subgroups brushed their teeth 0 to 1 times daily and always or mostly used dental floss and interdental brushes weekly, compared to the poorest oral health care group (*P*_trend_=0.037). A reduced risk of total stroke was also shown in the group with higher household income when they brushed their teeth at least twice daily, regardless of the frequency of weekly dental flossing and interdental brushing: those who never used dental floss and interdental brushes (aHR 0.84, 95% CI 0.70–1.00), those who sometimes used them (aHR 0.81, 95% CI 0.67–0.98), and those who always or mostly used them (aHR 0.82, 95% CI 0.68–0.99). However, significant within-group differences were not observed (all *P*interaction>0.05).

**Table 3.** Stratified analysis of risk for total stroke^a^ according to the frequency of toothbrushing, dental flossing and interdental brushing.

| Frequency of toothbrushing (times/day) | Never or once |  |  | Twice or more |  |  | p for trend | p for interaction |
| --- | --- | --- | --- | --- | --- | --- | --- | --- |
| Frequency of dental flossing and interdental brushing (frequency/week) | Never | Sometimes | Always or mostly | Never | Sometimes | Always or mostly |  |  |
| Number of participants (%) | 4,048 (4.09) | 1,251 (1.27) | 1,103 (1.12) | 47,330 (47.87) | 22,477 (22.73) | 22,657 (22.92) |  |  |
| **Total stroke<sup>a</sup>** |  |  |  |  |  |  |  |  |
| **Age** |  |  |  |  |  |  |  | 0.594 |
| **<65 years** |  |  |  |  |  |  |  |  |
| Events, n | 132 | 47 | 46 | 1,338 | 611 | 630 |  |  |
| Person-years | 1,026 | 394 | 437 | 11,231 | 5,387 | 5,685 |  |  |
| aHR (95% CI) | **1.00 (Ref.)** | 0.89 (0.63-1.24) | 0.76 (0.55-1.07) | 0.85 (0.71-1.02) | 0.83 (0.69-1.01) | **0.78 (0.65-0.95)** | 0.016 |  |
| **≥65 years** |  |  |  |  |  |  |  |  |
| Events, n | 124 | 24 | 13 | 627 | 138 | 223 |  |  |
| Person-years | 885 | 179 | 113 | 4,225 | 1,022 | 1,483 |  |  |
| aHR (95% CI) | **1.00 (Ref.)** | 1.22 (0.79-1.90) | 0.71 (0.40-1.29) | 1.12 (0.92-1.36) | 1.02 (0.80-1.31) | 1.13 (0.90-1.42) | 0.390 |  |
| **Sex** |  |  |  |  |  |  |  | 0.353 |
| **Male** |  |  |  |  |  |  |  |  |
| Events, n | 180 | 60 | 41 | 1,088 | 462 | 529 |  |  |
| Person-years | 1,382 | 485 | 409 | 9,416 | 4,362 | 4,928 |  |  |
| aHR (95% CI) | **1.00 (Ref.)** | 1.04 (0.77-1.39) | 0.73 (0.52-1.03) | 0.94 (0.80-1.10) | 0.91 (0.76-1.08) | 0.85 (0.72-1.01) | 0.045 |  |
| **Female** |  |  |  |  |  |  |  |  |
| Events, n | 76 | 11 | 18 | 877 | 287 | 324 |  |  |
| Person-years | 529 | 88 | 141 | 6,039 | 2,047 | 2,240 |  |  |
| aHR (95% CI) | **1.00 (Ref.)** | 0.90 (0.48-1.70) | 0.98 (0.59-1.66) | 1.04 (0.82-1.33) | 1.01 (0.78-1.31) | 1.05 (0.81-1.35) | 0.799 |  |
| **Household income** |  |  |  |  |  |  |  | 0.506 |
| **Higher** |  |  |  |  |  |  |  |  |
| Events, n | 147 | 36 | 31 | 1,126 | 487 | 484 |  |  |
| Person-years | 1,011 | 326 | 290 | 9,155 | 4,300 | 4,201 |  |  |
| aHR (95% CI) | **1.00 (Ref.)** | 0.86 (0.60-1.24) | 0.79 (0.54-1.16) | **0.84 (0.70-1.00)** | **0.81 (0.67-0.98)** | **0.82 (0.68-0.99)** | 0.077 |  |
| **Lower** |  |  |  |  |  |  |  |  |
| Events, n | 109 | 35 | 28 | 839 | 262 | 369 |  |  |
| Person-years | 900 | 247 | 260 | 6,300 | 2,109 | 2,968 |  |  |
| aHR (95% CI) | **1.00 (Ref.)** | 1.30 (0.88-1.90) | 0.87 (0.57-1.33) | 1.16 (0.95-1.43) | 1.17 (0.93-1.48) | 1.07 (0.86-1.33) | 0.891 |  |
| **Smoking status** |  |  |  |  |  |  |  | 0.302 |
| **Never** |  |  |  |  |  |  |  |  |
| Events, n | 131 | 30 | 28 | 1,197 | 406 | 514 |  |  |
| Person-years | **921** | 229 | 253 | 8,818 | 3,232 | 3,761 |  |  |
| aHR (95% CI) | **1.00 (Ref.)** | 1.06 (0.71-1.58) | 0.92 (0.61-1.39) | 1.00 (0.83-1.20) | 0.97 (0.79-1.19) | 1.04 (0.85-1.26) | 0.698 |  |
| **Ever** |  |  |  |  |  |  |  |  |
| Events, n | 125 | 41 | 31 | 768 | 343 | 339 |  |  |
| Person-years | 990 | 344 | 297 | 6,637 | 3,177 | 3,407 |  |  |
| aHR (95% CI) | **1.00 (Ref.)** | 0.98 (0.69-1.40) | 0.73 (0.49-1.09) | 0.94 (0.78-1.14) | 0.91 (0.74-1.12) | **0.79 (0.64-0.97)** | 0.015 |  |
| **Alcohol consumption** |  |  |  |  |  |  |  | 0.683 |
| **No** |  |  |  |  |  |  |  |  |
| Events, n | 140 | 28 | 31 | 1,177 | 393 | 497 |  |  |
| Person-years | 1,036 | 234 | 308 | 8,835 | 3,172 | 3,941 |  |  |
| aHR (95% CI) | **1.00** | 1.02 | 0.77 | 1.01 | 0.97 | 0.99 | 0.859 |  |
|  | (Ref.) | (0.68-1.53) | (0.52-1.13) | (0.85-1.21) | (0.79-1.18) | (0.82-1.20) |  |  |
| **Yes** |  |  |  |  |  |  |  |  |
| Events, n | 116 | 43 | 28 | 788 | 356 | 356 |  |  |
| Person-years | 875 | 339 | 242 | 6,620 | 3,237 | 3,227 |  |  |
| aHR (95% CI) | **1.00**<br>(Ref.) | 1.02<br>(0.72-1.45) | 0.89<br>(0.59-1.34) | 0.94<br>(0.77-1.14) | 0.91<br>(0.73-1.13) | 0.85<br>(0.69-1.05) | 0.078 |  |
| **Physical activity** |  |  |  |  |  |  |  | 0.687 |
| **No** |  |  |  |  |  |  |  |  |
| Events, n | 148 | 38 | 33 | 1,007 | 297 | 345 |  |  |
| Person-years | 1,128 | 297 | 314 | 7,443 | 2,321 | 2,724 |  |  |
| aHR (95% CI) | **1.00**<br>(Ref.) | 1.07<br>(0.75-1.54) | 0.81<br>(0.55-1.19) | 1.07<br>(0.90-1.28) | 1.09<br>(0.89-1.33) | 1.01<br>(0.83-1.23) | 0.873 |  |
| **Yes** |  |  |  |  |  |  |  |  |
| Events, n | 108 | 33 | 26 | 958 | 452 | 508 |  |  |
| Person-years | 783 | 276 | 236 | 8,012 | 4,088 | 4,444 |  |  |
| aHR (95% CI) | **1.00**<br>(Ref.) | 0.99<br>(0.67-1.46) | 0.81<br>(0.53-1.25) | 0.86<br>(0.70-1.05) | 0.82<br>(0.66-1.01) | 0.82<br>(0.67-1.02) | 0.058 |  |
| **Missing teeth** |  |  |  |  |  |  |  | 0.225 |
| **No** |  |  |  |  |  |  |  |  |
| Events, n | 159 | 43 | 40 | 1,349 | 524 | 643 |  |  |
| Person-years | 1,262 | 390 | 377 | 10,720 | 4,581 | 5,353 |  |  |
| aHR (95% CI) | **1.00**<br>(Ref.) | 0.95<br>(0.68-1.34) | 0.98<br>(0.69-1.39) | 1.03<br>(0.87-1.22) | 1.00<br>(0.83-1.20) | 1.00<br>(0.84-1.19) | 0.810 |  |
| **Yes** |  |  |  |  |  |  |  |  |
| Events, n | 97 | 28 | 19 | 616 | 225 | 210 |  |  |
| Person-years | 649 | 183 | 173 | 4,735 | 1,828 | 1,815 |  |  |
| aHR (95% CI) | **1.00**<br>(Ref.) | 1.18<br>(0.77-1.82) | **0.59**<br>**(0.36-0.98)** | 0.85<br>(0.68-1.06) | 0.86<br>(0.67-1.10) | **0.77**<br>**(0.60-0.99)** | 0.037 |  |
| **Charlson comorbidity index** |  |  |  |  |  |  |  | 0.778 |
| **≤1** |  |  |  |  |  |  |  |  |
| Events, n | 66 | 22 | 13 | 548 | 236 | 247 |  |  |
| Person-years | 498 | 210 | 143 | 4,493 | 2,069 | 2,069 |  |  |
| aHR (95% CI) | **1.00**<br>(Ref.) | 0.87<br>(0.53-1.42) | 0.89<br>(0.49-1.63) | 1.00<br>(0.77-1.30) | 0.96<br>(0.73-1.28) | 0.94<br>(0.71-1.24) | 0.625 |  |
| **≥2** |  |  |  |  |  |  |  |  |
| Events, n | 190 | 49 | 46 | 1,417 | 513 | 606 |  |  |
| Person-years | 1,413 | 363 | 407 | 10,962 | 4,339 | 5,099 |  |  |
| aHR (95% CI) | **1.00**<br>(Ref.) | 1.10<br>(0.80-1.51) | 0.78<br>(0.56-1.08) | 0.95<br>(0.81-1.11) | 0.93<br>(0.78-1.10) | 0.90<br>(0.76-1.07) | 0.176 |  |

Never or once: brushed teeth 0-1 times per day. Twice or more: brushed teeth 2 or more times per day Never: never used dental floss and interdental brushes per week. Sometimes: occasionally used dental floss and interdental brushes per week. Always or mostly: always or mostly used dental floss and interdental brushes per week
Abbreviation: aHR= adjusted hazard ratio; CI= confidence interval
Cox proportional hazard model adjusted for age, sex, body mass index, systolic blood pressure, diastolic blood pressure, fasting serum glucose, total cholesterol, household income, smoking status, alcohol consumption, physical activity, missing teeth, and the Charlson Comorbidity Index.
<sup>a</sup>Defined as subjects who had experienced at least two days of hospitalization with I60–I69

Supplementary Table 1 describes the results of the stratified analysis for ischemic stroke risk, with the poorest oral health care group designated as the reference. Compared to this group, individuals who brushed their teeth at least twice daily and always or mostly used dental floss and interdental brushes weekly showed significantly reduced ischemic stroke risk across several subgroups: those under 65 years of age (aHR 0.65, 95% CI 0.50–0.86), males (aHR 0.76, 95% CI 0.61–0.96), higher household income (aHR 0.75, 95% CI 0.57–0.97), those who have ever smoked (aHR 0.66, 95% CI 0.50–0.87), those who do physical activity (aHR 0.71, 95% CI 0.53–0.95), those with missing teeth (aHR 0.60, 95% CI 0.43–0.83), and those with the CCI of 2 or more (aHR 0.76, 95% CI 0.60–0.96) (all *P*_trend_<0.05). Moreover, a significantly reduced risk was identified among individuals under 65 years of age who brushed their teeth at least twice daily and never used dental floss and interdental brushes weekly (aHR 0.77, 95% CI 0.59–0.96), and among those with higher household income who brushed their teeth at least twice daily and sometimes used dental floss and interdental brushes weekly (aHR 0.73, 95% CI 0.56–0.95), both compared to the poorest oral health care group. Compared to the poorest oral health care group, individuals with missing teeth showed a significantly lower risk of ischemic stroke in both those who brushed their teeth 0 to 1 times daily and always or mostly used dental floss and interdental brushes weekly (aHR 0.37, 95% CI 0.17–0.81), and those who brushed at least twice daily and never used dental floss and interdental brushes weekly (aHR 0.74, 95% CI 0.56–0.98). However, no significant within-group differences were identified (all *P*_interaction_>0.05).

## 4. Discussion

To the best of our knowledge, we are the first to globally investigate whether the frequency of dental flossing and interdental brushing provides additional benefits for stroke prevention beyond the impacts of increased toothbrushing frequency. Our findings indicate that those who brush their teeth at least twice daily and always or mostly used dental floss and interdental brushes weekly may have greater protection against stroke compared to the poorest oral health care group, significantly reducing the risk of ischemic stroke by up to 23%. In the stratified analysis, simply increasing the frequency of dental flossing and interdental brushing was associated with a lower risk of ischemic stroke among individuals with missing teeth who brushed their teeth 0 to 1 times daily. These results suggest that dental flossing and interdental brushing alone may reduce the risk of stroke.

Biological mechanisms have been advanced to explain the link between oral hygiene practices, such as toothbrushing, dental flossing, and interdental brushing, and stroke. Poor oral hygiene may result in systemic inflammation and local infections, which are important risk factors for stroke, due to the gram-negative bacterial load and the subgingival biofilm in the mouth^25,26^. Participants who self-reported never or rarely brushing their teeth showed increased concentrations of C-reactive protein, an inflammatory marker^27^. Furthermore, periodontal bacteria and their toxic byproducts can enter the bloodstream through ulcerated periodontal epithelial tissue^28,29^. This may trigger an increased risk of atheromatous plaque formation or rupture, potentially leading to stroke^30^. In murine models, accelerated atherosclerosis has been attributed to *P. gingivalis*^31^.

Previous studies consistently have reported that good oral hygiene behaviors, including frequent toothbrushing and the use of interdental cleaning devices, can reduce the risk of cardiovascular disease. Toothbrushing, dental flossing, and interdental brushing all share the benefit of removing dental plaque. Chang et al. identified that brushing teeth 3 or more times daily can decrease the risk of stroke^25^. However, toothbrushing alone is insufficient to completely remove dental plaque from interdental areas^14,16^. Thus, the use of interdental cleaning methods is imperative, as they effectively remove more dental plaque, including areas that toothbrushes cannot reach^15^. The American Dental Association states that dental flossing can eliminate a maximum of 80% of plaque^32^. Schmid et al. found that the additional use of interdental toothbrushes can lead to a plaque removal effect of up to 96%^33^. According to Lee et al., using dental floss or interdental toothbrushes is linked with a 16% lower risk of cardiovascular and cerebrovascular diseases^34^. This result corresponds to our findings, which suggest that the impact on reduced stroke risk may be enhanced by the additional use of interdental cleaners, as well as the benefits of regular toothbrushing.

Among these studies, the research by Park et al. stands out as the first to investigate the relationship between oral hygiene practices, including tooth brushing, and cardiovascular disease^4^. They used a baseline population of individuals who were subject to health examinations from 2002 to 2003 and tracked these participants until 2013^4^. The outcome was not only the occurrence of stroke but also heart failure and myocardial infarction^4^. In comparison to the study performed by Park et al., our study adopts a different approach by broadening the scope of oral hygiene to include dental flossing and interdental brushing as well as toothbrushing. Moreover, we focused exclusively on stroke as the outcome, allowing us to conduct a detailed analysis of stroke risk in relation to the use of dental floss and interdental brushes. Additionally, our study is according to a more recent baseline population, consisting of individuals who underwent health examinations in 2009-2010, with follow-up until 2019, enabling us to assess more contemporary trends. This distinction underscores the originality and relevance of our research within the current landscape.

In our stratified analysis, the alcohol consumption variable showed no statistical significance. On the other hand, we observed a consistent trend toward a reduced risk of both total stroke and ischemic stroke among those under 65 years of age, those with higher household income, those who have ever smoked, and those with missing teeth who increased their frequency of toothbrushing, dental flossing, and interdental brushing. Particularly in the group with missing teeth, always or mostly using dental floss and interdental brushes was related to a significantly reduced risk of total stroke and ischemic stroke, regardless of daily toothbrushing frequency. This can be interpreted as individuals with missing teeth may experience a greater reduction in stroke risk even with slight improvements in oral hygiene practices, likely due to their poor oral health. Therefore, the importance of using interdental cleaners such as dental floss and interdental brushes cannot be overemphasized.

There are several limitations to our study. First, the evaluation of oral hygiene practices frequency, such as toothbrushing, dental flossing, and interdental brushing, may be inaccurate due to the basis of self-reported questionnaires. Second, this analysis assumed that oral care frequencies remained constant after follow-up began, despite potential changes in oral hygiene habits over time. Moreover, the absence of statistical significance for hemorrhagic stroke is likely due to the small sample size. Finally, the NHIS database lacked information on factors such as the number of remaining teeth, the use of other interdental cleaners, exclusive use of either dental floss or interdental brushes, and data on younger participants in their twenties. Previous studies have discussed the impact of the number of remaining teeth on the efficacy of interdental cleaners^15,35^. The NHIS database began including data on dental flossing and interdental brushing frequency, the exposure variables in this study, in 2009. However, it only provides information on the presence of missing teeth, not their number. In addition, the database did not include details on the use of other interdental cleaning devices or exclusive use of a single type^15,34,36–38^. Although previous research indicated the effectiveness of using dental floss and interdental brushes among younger subjects^15^, our study could only include subjects aged 40 years or older due to the characteristics of database. Future research with more comprehensive data is essential to address the limitations of our study.

Our study, notwithstanding these limitations, demonstrates several strengths. It is the first to examine the nationwide population aged 40 years or older in Korea, using the NHIS database from 2009 onwards, when data on dental flossing and interdental brushing was initially collected. VanWormer et al. assessed the relationship between oral hygiene practices and cardiovascular diseases risk^39^ but could not establish clear causality due to their cross-sectional study design. In contrast, our retrospective cohort study identified that using a toothbrush, dental floss, and interdental brushes separately may reduce stroke risk.

While there is the widespread belief that increased toothbrushing contributes to better health, some studies proposed that excessive toothbrushing might lead to negative consequences such as tooth wear^40^ and gingival recession^41,42^. Despite these controversies, our study supports the positive impacts of toothbrushing on health. Furthermore, the validity of our findings was strengthened by considerable adjustment for various confounders in the stratified analysis. Education level, which can influence both dental flossing^43,44^ and cardiovascular diseases^45–48^, may have served as a confounder in this study. Since the NHIS data lack participants’ educational information^49^, household income may act as a proxy.

While toothbrushing is widely practiced due to well-publicized policies and campaigns^50^, the benefits of dental flossing and interdental brushing remain relatively unknown, with limited policy support and campaigns encouraging their use. Thus, it is important to raise awareness of these additional benefits through policies and campaigns, utilizing our paper as a reference. Public education on effective self-performed oral hygiene practices is also essential^51^. Therefore, establishing specific guidelines not only for toothbrushing but also for the proper use of dental floss and interdental brushes is necessary.

In conclusion, this study specifically proposes a link between frequent dental flossing and interdental brushing and a lower risk of stroke. We underscore the importance of good oral hygiene practices as a helpful strategy for stroke prevention. Our findings indicate that enhancing oral hygiene practices can significantly lower stroke risk in individuals with poor oral health. Further research is needed to verify these results and draw more definitive conclusions. Since dental floss and interdental brushes may be readily available, incorporating these methods would be advantageous for both reducing stroke risk and improving oral health.

## Ethical Statement

This study received thorough review and approbation from the Institutional Review Board of Seoul National University Hospital (IRB number: E-2311-054-1483). Since the NHIS data used for analysis was anonymized in accordance with stringent confidentiality guidelines prior to its release, obtaining informed consent was considered unnecessary for this study.

## CRediT authorship contribution statement

Da Eun Kim, Sangwoo Park, and Sang Min Park were involved in the conception and design, data analysis and interpretation, critical review for significant intellectual content, and gave final approval for the article. Da Eun Kim and Sangwoo Park contributed equally to drafting the article and are co-first authors. Da Eun Kim collected and organized the data. The final version of the manuscript was approved by all authors. Sang Min Park, as the corresponding author, had full access to all data in the study and assumes responsibility for the data’s integrity and the precision of the analysis.

## Declaration of Competing Interest

The authors declare that there are no known financial conflicts of interest, activities, relationships, or affiliations that could have influenced the work reported in this manuscript. All potential conflicts of interest have been disclosed in full compliance with the journal’s requirements, including specific financial interests relevant to the subject of the manuscript. The information provided is accurate and reflects the most recent status according to the guidelines provided.

## Funding

This research was funded by the Seoul National University Hospital Research Fund and received support from the BK21 FOUR education program scholarship provided by the National Research Foundation of Korea.

## Data availability

Data will be provided upon reasonable request. For data requests, please contact the corresponding author for further details.

---

**Supplementary Table 1.**
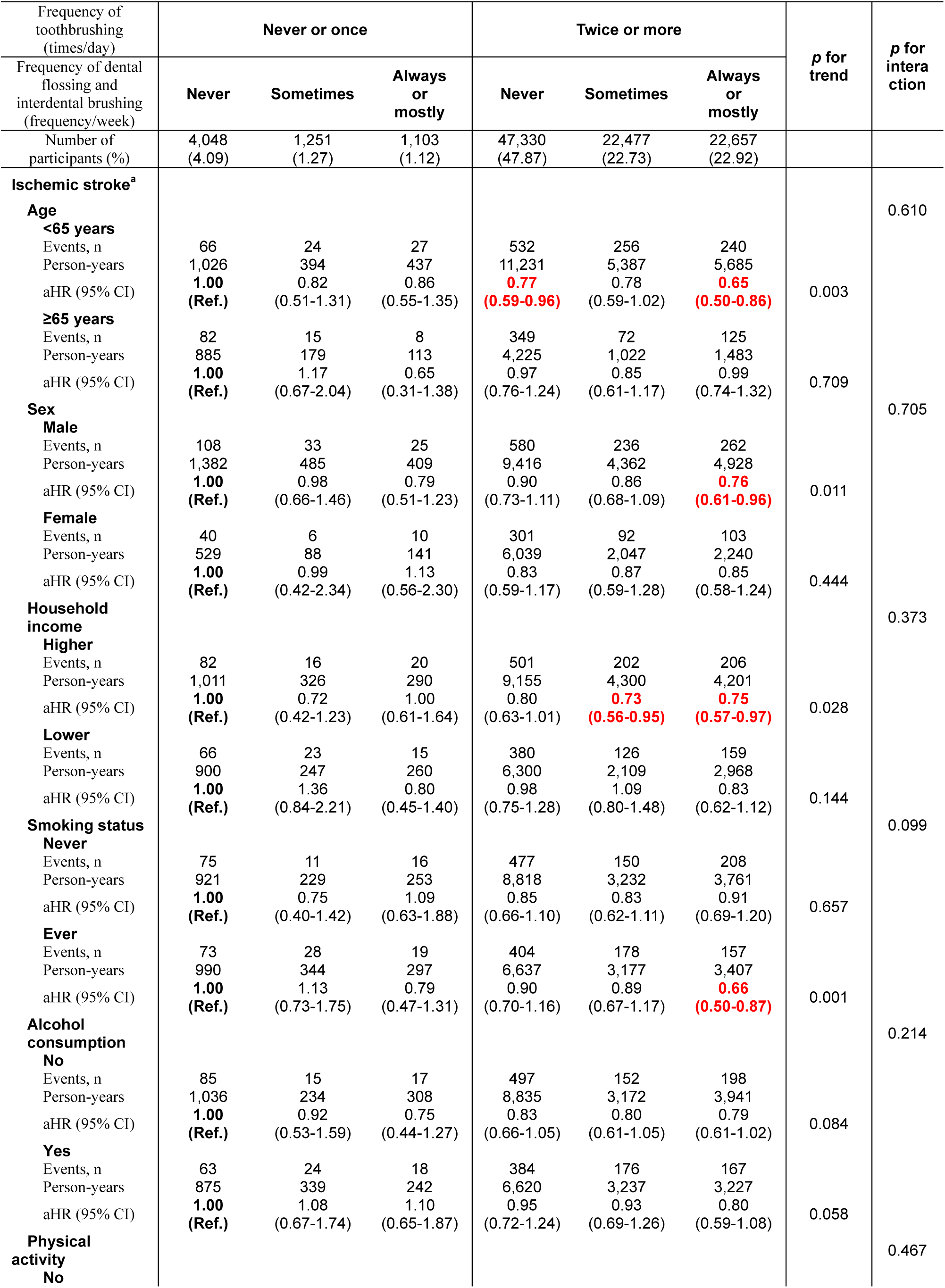
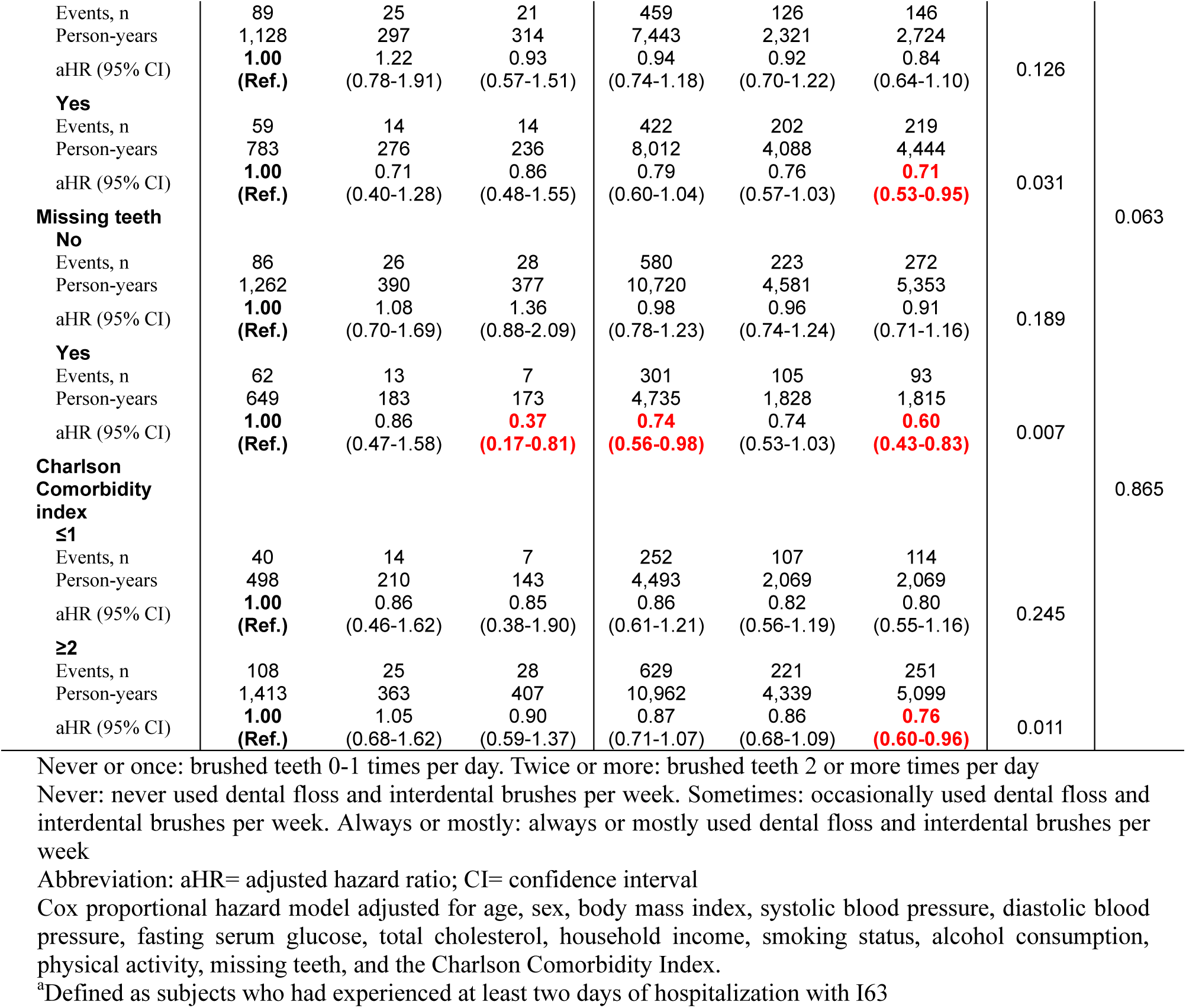
Stratified analysis of risk for ischemic stroke^a^ according to the frequency of toothbrushing, dental flossing and interdental brushing.

## Notes

### Competing Interest Statement

The authors have declared no competing interest.

